# Early Intervention in preterm infants modulates LINE-1 promoter methylation and neurodevelopment

**DOI:** 10.1101/19011874

**Authors:** Camilla Fontana, Federica Marasca, Livia Provitera, Sara Mancinelli, Nicola Pesenti, Shruti Sinha, Sofia Passera, Sergio Abrignani, Fabio Mosca, Simona Lodato, Beatrice Bodega, Monica Fumagalli

## Abstract

**Background:** Early life adversity exposure impacts preterm infants’ neurodevelopment and early intervention protocols may modulate neurodevelopmental outcomes.

Neuronal genomes are plastic in response to environment and mobile genetic elements, including LINE-1 (L1), are source of brain genomic mosaicism. Maternal care during early life regulates L1 methylation and copy number variations (CNVs) in mice. Here, we sought to identify the effects of maternal care and positive multisensory stimulation (Early Intervention) on L1 methylation and neurodevelopment in preterm infants.

**Methods:** Very preterm infants were randomized to receive Standard Care or Early Intervention. L1 methylation was measured at birth and at hospital discharge. At 12 months infants’ neurodevelopment was evaluated with the Griffiths Scales. L1 methylation and CNVs were measured in mouse brain areas at embryonic and postnatal stages.

**Results:** We demonstrated that L1 is hypomethylated in preterm versus term infants at birth. Early Intervention contributes to restore L1 methylation and positively modulates neurodevelopment. We showed that L1 methylation is developmentally-regulated in mice, decreasing in early postnatal life stages, which turns into an increased L1 CNVs specifically in hippocampus and cortex.

**Conclusions:** Here we demonstrated that L1 dynamics can be modulated by Early Intervention, in parallel with ameliorated neurodevelopmental outcomes. We further identified a specific developmental window of the fetal mouse brain, sensitive to early life experience, in which L1 dynamics are fine-tuned contributing to shape the brain genomic landscape.

**Trail Registration:** clinicalTrial.gov (NCT02983513)

**Funding:** Italian Ministry of Health (RC 780/03 2017), University of Milan (DISCCO 2015) and INGM internal funding.

## INTRODUCTION

Prematurity, which is defined as birth before 37 weeks of gestation, affects 11% of neonates globally, it is the second leading cause of death in children below 5 years of age and the most important in the first month of life (1). Among preterm infants, about 16% are born very preterm (<32 weeks of gestation) (2). This condition is associated with a considerable risk to develop acute and chronic postnatal morbidities and long-term neurodevelopmental disabilities (3). Indeed, up to 15% of very preterm infants suffer from severe neurologic disorders, mainly related to the occurrence of acquired brain lesions (4) and up to 50% of preterms experience other neurocognitive impairments in different areas of development (e.g. language, behavior, visual processing, academic performances and executive functions) (5, 6) or neuropsychological problems, including Attention Deficit/Hyperactivity Disorder (ADHD) or Autism Spectrum Disorders (ASD) (7-10).

Neurodevelopmental delays may occur even in absence of overt acquired brain lesions and are most likely caused by prematurity - related impairment in brain microstructural maturation and connectivity (11-13). Besides the documented role played by gestational age (GA) at birth and by the occurrence and severity of postnatal morbidities, the impact of early exposure to the hazards of extrauterine life has been recently emphasized. Indeed, during their stay in Neonatal Intensive Care Unit (NICU) preterm infants face early environmental stress, mainly represented by the excessive neurosensory stimulation and the prolonged separation from their parents (14, 15). Overall, these environmental stressors act in a critical window of the preterm brain development (corresponding to the last trimester of pregnancy and early postnatal life), in which multiple biological processes are taking place, including the earliest myelination phase, neuronal circuit assembly, and synaptogenesis (16). Notably, there is a 4-fold increase in cerebral cortical volume during the last trimester, which is accompanied by a significant increase in brain surface area, resulting in the formation of sulci and gyri of the cerebral cortex (17). Cognitive and emotional processing relays on the proper development of the cerebral cortex (which includes both the neocortex and the hippocampus) (18, 19). Alteration of the finely orchestrated and precisely timed development of cortical and hippocampal neural circuits has been associated to prominent long-term consequences in childhood and adulthood (20-22).

Interestingly, recent findings have highlighted the crucial role of parental care on modulating the detrimental effects of early life exposure (23). Therefore, Developmental Care has been conceived as a strategy to reduce NICU stressful factors and promote maternal engagement; it has been demonstrated to improve brain maturation, as assessed by magnetic resonance imaging (MRI), and neurodevelopmental outcomes (24-26). More recently, Early Intervention strategies, based on a multisensory stimulation approach, have been shown to promote infants’ visual acuity, neurobehavior and brain development (27-29).

How these early life experiences can modulate at molecular level the brain architecture and, ultimately, child’s behavior is still an unsolved issue. Interestingly, it has been recently demonstrated that neuronal genomes are plastic in response to environmental cues; in particular, early maternal care can affect genome structural variations in mouse hippocampus, thus resulting in somatic mosaicism that ultimately generates neuronal diversity with potential effect on behavior (30-34).

Mobile DNA elements have the ability to change their genomic position, either by a DNA-based (transposition) or RNA-based (retrotransposition) mechanism. Retrotransposition is one of the main forms of somatic mosaicism in the brain (35). Among retrotransposable elements, LINE-1 (L1), that covers about 18% of the human genome (36), has been extensively described to retrotranspose in neurons from fly to humans (37-39), a mechanism that takes place during neural progenitor development and differentiation (40-42). L1 can move to different genomic location (*de novo* insertion site) by the activity of a reverse transcriptase (RTase), encoded by L1 itself, which reverse-transcribes and integrates a L1 cDNA copy, that is usually 5’ end truncated (36).

L1 activity is finely modulated (42, 43) at the level of its endogenous promoter, where a CpG island methylation/demethylation is associated with L1 somatic mobilization in the brain (43). Notably, maternal care during early life has been reported to drive variability in L1 methylation and copy number variations (CNVs) within the mouse hippocampal neuronal genome, influencing progeny behavior (31). Similarly, childhood stress and adversities in early life have been reported to result in L1 hypomethylation (44, 45). Furthermore, the deregulation of L1 activity has been described in the brain of debilitating neurological diseases as Rett syndrome (43, 46), schizophrenia (47, 48), autism (49), bipolar and major depressive disorder (33, 50).

In the current study we combined analyses in humans and mice to test the hypothesis that an early intervention program in very preterm infants, based on maternal care and enriched multisensory stimulation, could modulate L1 dynamics in the developing brain and have a clinical readout on short-term neurodevelopment.

## RESULTS

### Characteristics of study participants

Overall, 70 very preterm infants born between 25^+0^ and 29^+6^ weeks of gestational age (GA) were recruited and randomized either to receive Standard Care or Early Intervention (Standard Care n = 36, Early Intervention n = 34) between April 2014 and January 2017 (Figure 1). Standard Care, in line with NICU routine care protocols, included Kangaroo Mother Care and minimal handling. Early Intervention, in addition to routine care, included a parental training program together with enriched multisensory stimulation (infant massage and visual interaction, see Methods) promoted by parents as fully described in (51). A daily diary was given to parents to record all the interventions performed and to retrospectively quantify the effects of maternal care and multisensory stimulation. The study was conducted in a NICU with open access to parents (24 hours a day for 7 days a week).

**Figure 1.**
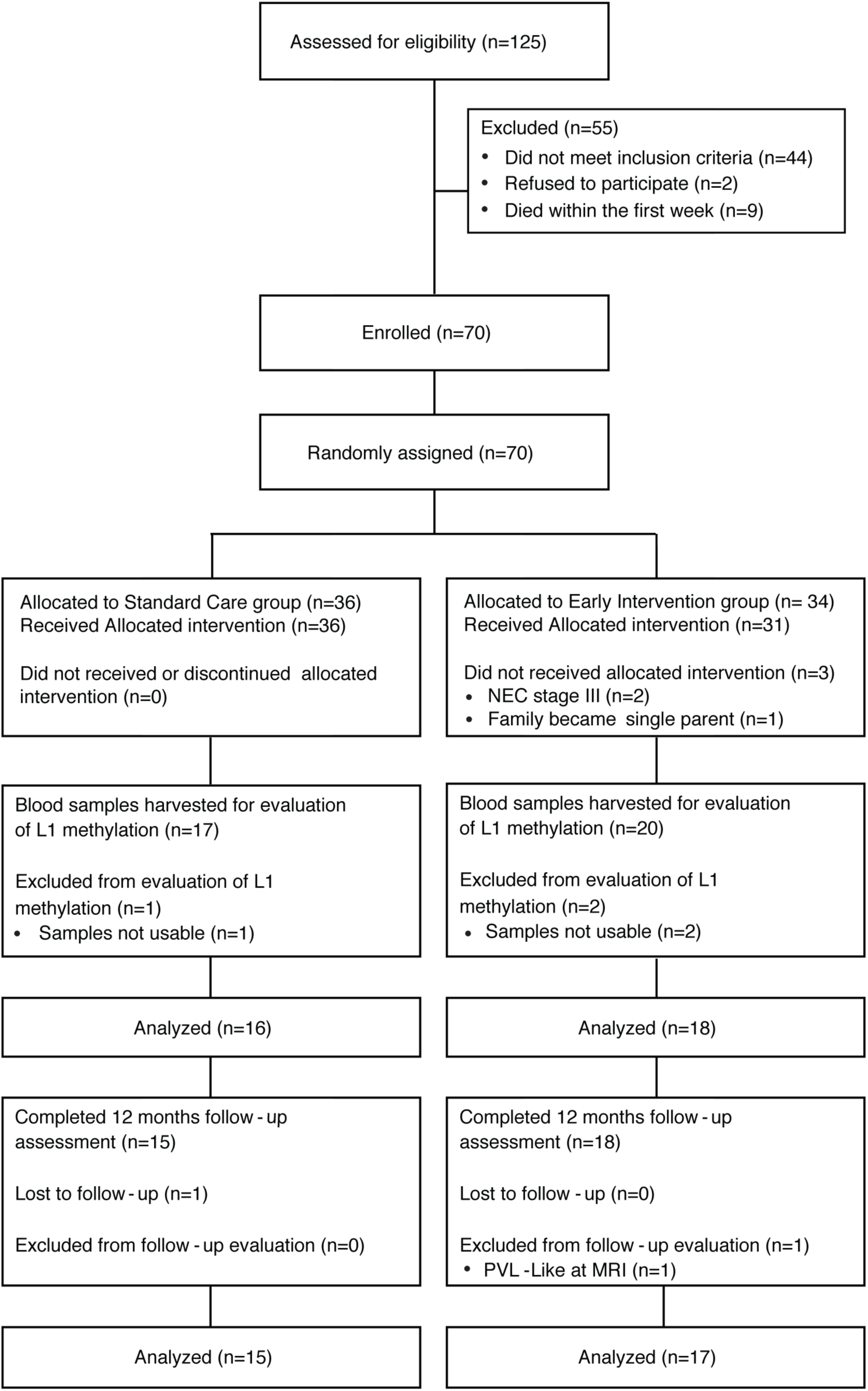
Flow chart of the study. CONSORT flow diagram showing patient enrollment, allocation to Standard Care and Early Intervention groups, subsequent L1 promoter methylation analysis and neurodevelopmental evaluation.

According to the protocol, 3 infants allocated to Early Intervention did not receive the treatment, as 2 infants developed surgical Necrotizing Enterocolitis (NEC) and 1 belonged to a family that became single-parent after the enrollment (Figure 1). All infants in the Standard Care group received allocated treatment as part of the routine clinical practice.

A schematic representation of the study timeline is provided in Figure 2. We harvested blood from 17 infants belonging to the Standard Care group and 20 to the Early Intervention group. Three infants (Standard Care n=1, Early Intervention n=2) were subsequently excluded as samples were not usable for molecular analyses (Figure 1). In details, L1 methylation analysis was carried out on 19 cord blood samples collected at birth, named as “preterm” and 33 peripheral blood samples collected at discharge, named as Standard Care (n=16) and Early Intervention (n=17). Baseline and perinatal characteristics for the two groups are described in Table 1; no differences were observed among the groups in terms of maternal and infant characteristics, or incidence of postnatal morbidities during NICU stay. Of note, infants enrolled in the study were discharged around Term Equivalent Age (TEA) with no significant differences between the two groups (Table 1).

**Table 1.** Baseline characteristics of the population: descriptive statistics and comparisons between Early Intervention and Standard Care groups

| Demographic feature | Standard Care<br>(n = 16) | Early<br>Intervention<br>(n = 18) | p value |
| --- | --- | --- | --- |
| **Maternal characteristics** |  |  |  |
| Maternal Age (years), mean± SD | 35.1 ± 6.1 | 33.1 ± 4.8 | 0.293<sup>^</sup> |
| Education (up to high school), n (%) | 8 (50%) | 3 (17%) | 0.066<sup>°</sup> |
| Socio-economic status, mean ± SD | 47.8 ± 15.8 | 51.3 ± 9.3 | 0.627<sup>*</sup> |
| Maternal smoking before or during pregnancy, n (%) | 2 (12%) | 1 (6%) | 0.591<sup>°</sup> |
| Maternal alcohol abuse during pregnancy, n (%) | 0 (0%) | 0 (0%) | >0.999<sup>°</sup> |
| Clinical chorioamnionitis, n (%) | 8 (50%) | 5 (28%) | 0.291<sup>°</sup> |
| **Infant characteristics** |  |  |  |
| Gestational age at birth (weeks), mean ± SD | 27.9 ± 1.1 | 28.1 ± 1.4 | 0.318<sup>*</sup> |
| Birth Weight (g), mean ± SD | 1089 ± 347 | 1005 ± 296 | 0.453<sup>^</sup> |
| Male, n (%) | 9 (56%) | 9 (50%) | 0.744<sup>°</sup> |
| Singleton, n (%) | 6 (38%) | 8 (44%) | 0.738<sup>°</sup> |
| Small for Gestational Age, n (%) | 3 (19%) | 4 (22%) | >0.999<sup>°</sup> |
| Cesarean Section, n (%) | 14 (88%) | 18 (100%) | 0.214<sup>°</sup> |
| Apgar score at 1', median (range) | 7 (2-8) | 6 (4-8) | 0.832<sup>*</sup> |
| Apgar score at 5', median (range) | 8 (5-9) | 8 (6-9) | 0.409<sup>*</sup> |
| Clinical Risk Index for Babies (CRIB) II score, mean ± SD | 8.0 ± 2.5 | 8.1 ± 2.1 | 0.903<sup>*</sup> |
| Days of Hospitalization, mean ± SD | 79.4 ± 30.7 | 83.9 ± 27.6 | 0.654<sup>^</sup> |
| Gestational Age at Discharge (weeks), mean ± SD | 39.4 ± 3.7 | 40.1 ± 3.7 | 0.567<sup>^</sup> |
| Days in the incubator, mean ± SD | 51.9 ± 21.9 | 55.7 ± 18.8 | 0.589<sup>^</sup> |
| **Postnatal Morbidities** |  |  |  |
| Days of Invasive Mechanical Ventilation, mean ± SD | 4.2 ± 6.3 | 6.1 ± 8.8 | 0.914<sup>*</sup> |
| Days of Non-Invasive Ventilation (NCPAP + nHFT), mean ± SD | 31.9 ± 20.4 | 49.1 ± 36.1 | 0.220<sup>*</sup> |
| Sepsis, n (%) | 5 (31%) | 11 (61%) | 0.101<sup>°</sup> |
| Severe Bronchopulmonary Displasia, n (%) | 2 (12%) | 8 (44%) | 0.063<sup>°</sup> |
| Germinal Matrix Hemorrhage - Intraventricular Hemorrhage (GMH - IVH) 1-2, n (%) | 2 (12%) | 2 (11%) | >0.999<sup>°</sup> |
| Retinopathy of Prematurity (ROP) any grade, n (%) | 2 (12%) | 2 (11%) | >0.999<sup>°</sup> |

Values are shown as count (percentage) for categorical variables and means ± standard deviations or median (range) for continuous variables. P-values were obtained using t-test (<sup>^</sup>), Mann-Whitney U Test (<sup>\*</sup>) or Fisher Exact Test (<sup>°</sup>).

**Figure 2.**
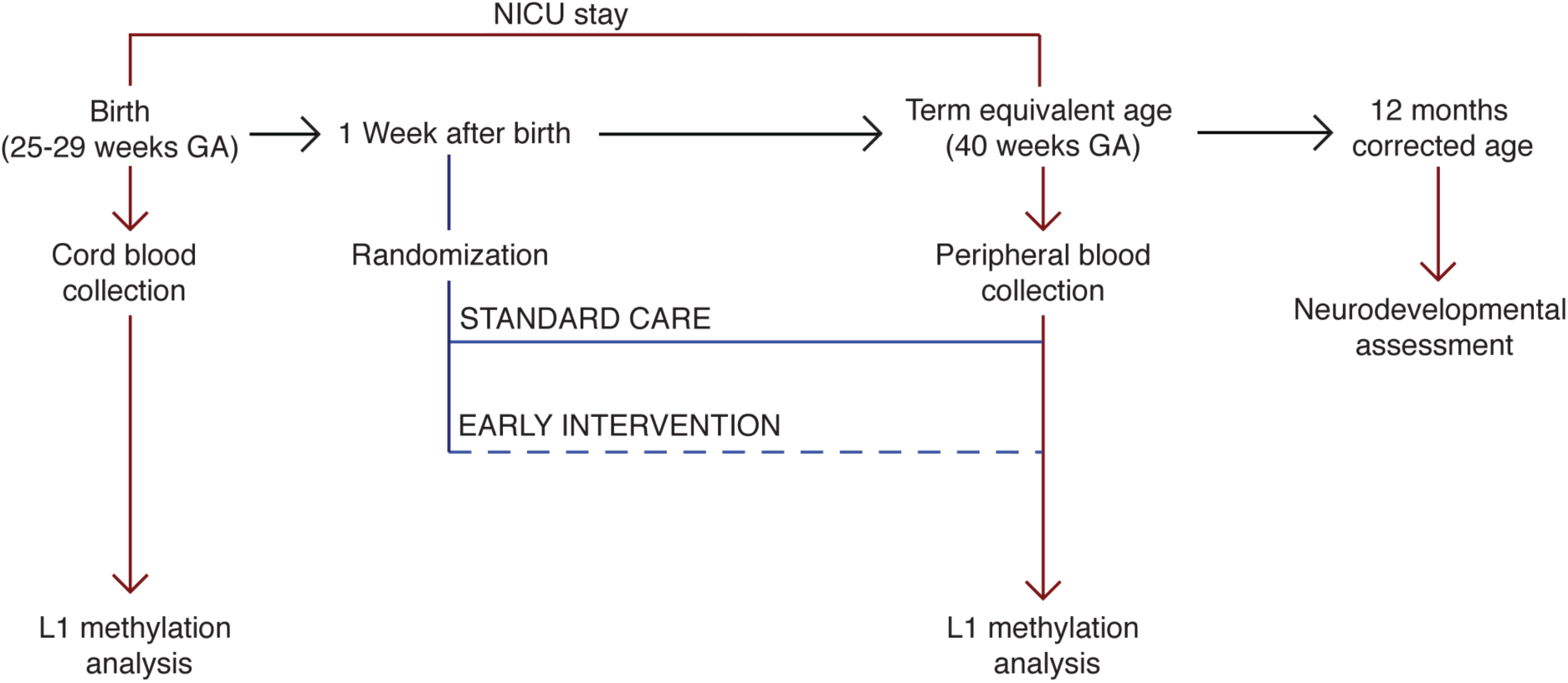
Time line of the study. Preterm infants born between 25^+0^ and 29^+6^ weeks gestational age (GA) were recruited. At birth, cord blood samples were collected. One week after birth, preterm infants were randomized to either receive Standard Care or Early Intervention during NICU stay. At term equivalent age (40 weeks GA), before NICU discharge, peripheral blood samples were harvested. At 12 months corrected age neurodevelopmental assessment was performed. L1 promoter methylation analysis was performed on genomic DNA extracted form cord blood and peripheral blood.

In the Early Intervention group the massage therapy was started by parents at 31.7 ± 1.8 (mean±SD) weeks of GA and carried out with a mean of 10.0 ± 2.1 times a week. Visual interaction was proposed from 34.9 ± 0.8 weeks of GA, and performed with a mean of 7.1 ± 1.8 times a week.

In addition, 21 cord blood samples from healthy full-term infants, named as “full-term”, were collected at birth (blood sample from 1 infant was excluded due to postnatal complications). Mother and infants’ characteristics of the full-term group were: mean maternal age at child birth 37.2 ± 3.8 years, mean GA at birth 38.5 ± 0.5 weeks, mean birth weight 3234 ± 420 g and median Apgar score 9 at 1 minute and 10 at 5 minutes; 30% were males.

### L1 promoter is hypomethylated in preterm infants at birth and its methylation level is restored upon Early Intervention

To assess L1 methylation level, we analyzed a region within L1 promoter, containing the CpG island already reported to modulate L1 transcription and activity (42); this CpG island is constituted of 19 CpG, of which CpG 11-19 are specifically involved in L1 regulation in the brain (Figure 3a, Neural specific CpG 11-19) (42); moreover, YY1 transcription factor binding site, required for neural specific L1 expression, corresponds to CpG 17 (52) (Figure 3a).

**Figure 3.**
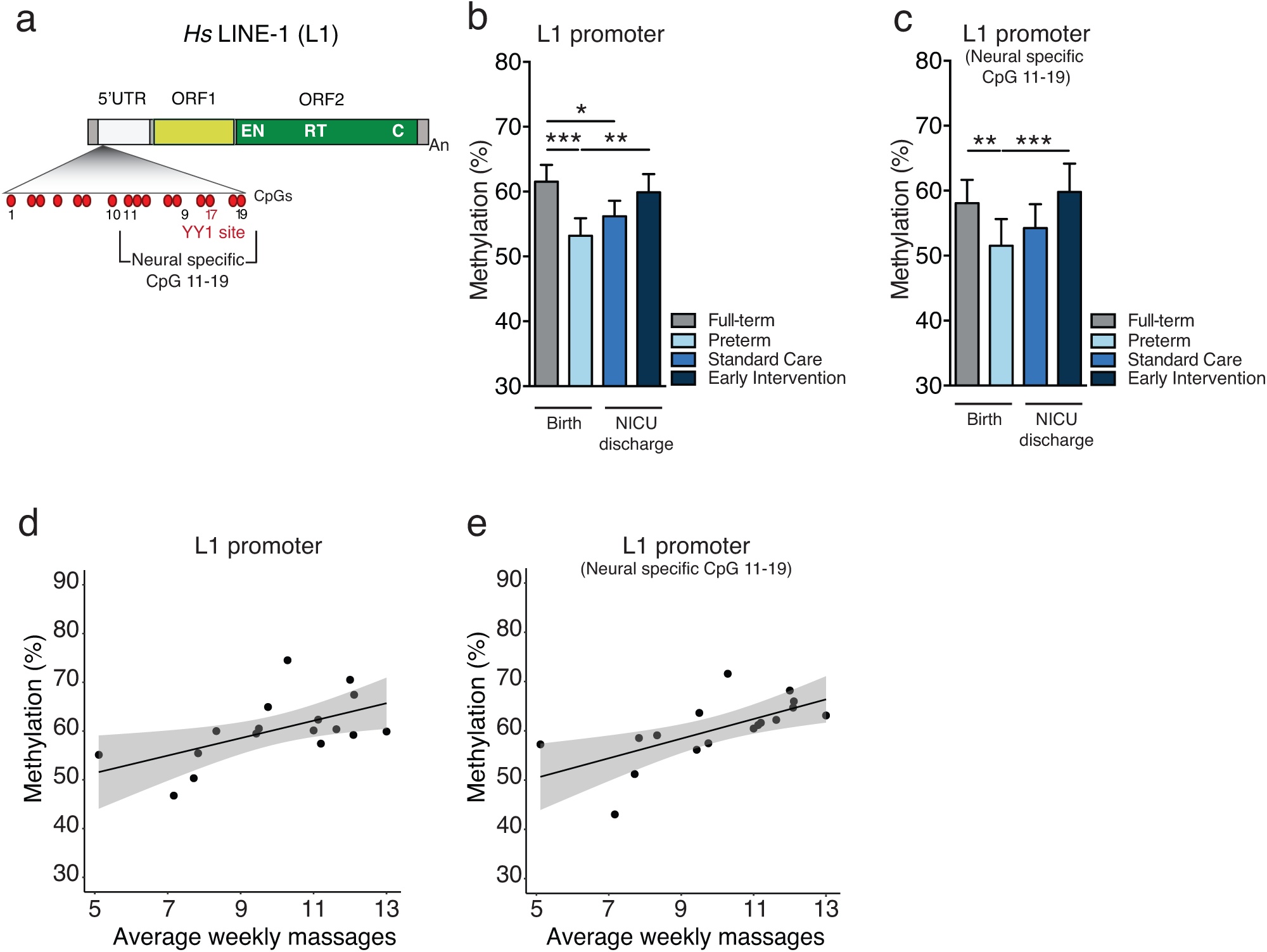
L1 promoter is hypomethylated in preterm neonates and its methylation is restored upon Early Intervention at NICU discharge. **a)** Schematic representation of human (*Hs*) LINE-1 (L1): 5’ untranslated region (5’UTR) that contains the internal promoter (arrow), open reading frame 1 (ORF1) and open reading frame 2 (ORF2). ORF2 includes endonuclease (EN), reverse transcriptase (RT), and cysteine-rich domains (C); poly (A) tract (An). Within the L1 promoter are highlighted: CpG island (CpG 1-19), the neural specific CpG (CpG 11-19, as reported in (42)) and YY1-binding site. **b-c)** Methylation analysis of **b)** L1 promoter and **c)** L1 promoter neural specific CpG 11-19 performed on genomic DNA extracted from cord blood of full-term (n = 20) and preterm neonates (n = 19) at birth and from peripheral blood of preterm infants at NICU discharge treated with Standard Care (n = 16) or Early Intervention (n = 17). In **b)** ***p < 0.001, Full-term vs Preterm; *p = 0.015, Full-term vs Standard Care; **p = 0.008, Preterm vs Early Intervention; in **c)** **p = 0.001, Full-term vs Preterm; ***p < 0.001, Preterm vs Early Intervention, unpaired two-tailed t test. **d-e)** Scatter plot and linear regression line with 95% confidence band of weekly infant massages vs **(d)** L1 promoter methylation level (estimate: 1.8, p = 0.017) and vs **(e)** L1 promoter neural specific CpG 11-19 methylation level (estimate: 2.0, p = 0.005) in the Early Intervention group (n = 17).

L1 promoter methylation level was analyzed as described in Coufal et al. 2009 (42) (see Methods) in the cord blood DNA of full-term (n=20) and preterm (n=19) neonates and in the peripheral blood DNA of preterm infants at NICU discharge, subjected either to Standard Care (n=16) or Early Intervention (n=17) (Figure 2 and Table 1). We found that L1 promoter methylation at birth was significantly lower in preterm compared to full-term cord blood (Figure 3b and Supplemental Figure 1a-b); at NICU discharge, L1 methylation levels were restored to level comparable to full-term infants only in the infants belonging to the Early Intervention group (Figure 3b and Supplemental Figure 1a-d). L1 methylation recovery in the Early Intervention group was specific for the neural region of the promoter (CpG 11-19) (Figure 3c and Supplemental Figure 1e) and in particular for CpG 17 corresponding to YY1 binding site (Supplemental Figure 1f).

Noteworthy, L1 promoter methylation level increases proportionally to the maternal care received upon Early Intervention (average number of massages received per week, recorded by parents in daily diary) (Figure 3d), a trend specifically observed for the neural region of L1 promoter (CpG 11-19) (Figure 3e) and not for CpG 1-10 (Supplemental Figure 1g). L1 methylation levels did not reflect a global genome methylation trend, as the promoter of a neuronal gene, *NAB2* (NGFI-A binding protein 2), did not show any difference in DNA methylation among the groups analyzed (Supplemental Figure 2a); moreover, the observed differences were unrelated to different cord blood cellular composition between full-term and preterm infants (Supplemental Figure 2b-c, see Methods).

### L1 promoter methylation is dynamically regulated during hippocampus and cortex devolvement in mice

To analyze L1 promoter methylation during brain development, we dissected presumptive somatosensory cortex, hippocampus and cerebellum from the mouse brain at multiple embryonic (E15.5 and E18.5), early postnatal (P0, P3) and later developmental stages (P14) (Figure 4a). We performed L1 methylation analysis as reported in Bedrosian et al (31) (see Methods) on L1MdTf family, the most active and evolutionary young L1 subfamily in mice (53); L1MdTf 5’UTR is constituted by several monomers, each containing a CpG island of 13 CpGs with a YY1 binding site corresponding to the CpG 8 and 9 (54) (Figure 4b).

**Figure 4.**
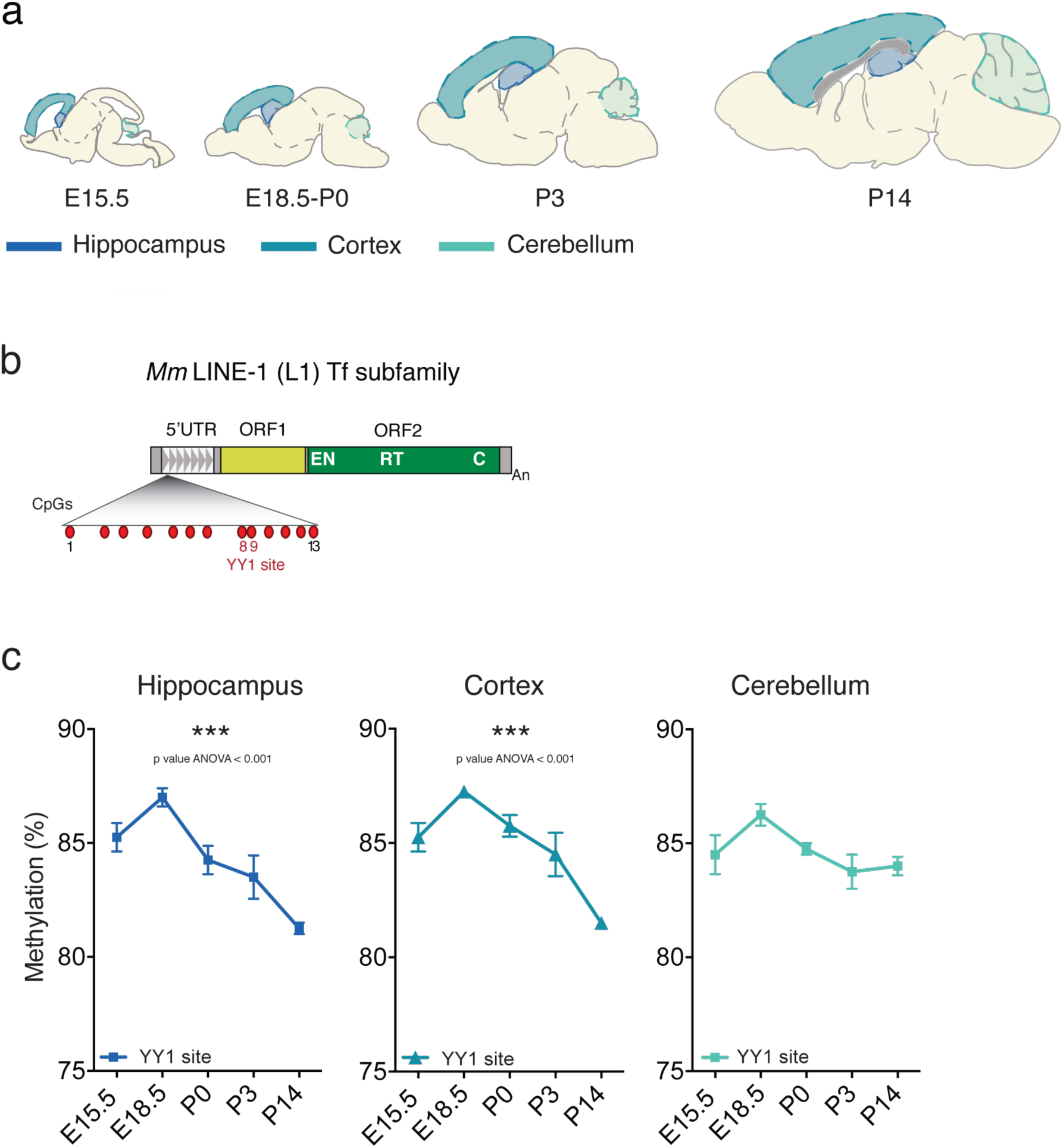
L1 promoter methylation levels are dynamic during development in mouse brain. **a)** Schematic drawings representing sagittal sections of mouse brain at different stages of embryonic-perinatal (E15.5, E18.5, P0) and postnatal (P3, P14) development. The regions micro-dissected for DNA methylation assay (hippocampus, cerebral cortex and cerebellum) are highlighted with different tones of blue. **b)** Schematic representation of mouse (*Mm*) LINE-1 (L1) Tf subfamily (L1MdTf): 5’ untranslated region (5’UTR), monomeric repeats (white triangles), open reading frame 1 (ORF1) and open reading frame 2 (ORF2). ORF2 includes endonuclease (EN), reverse transcriptase (RT), and cysteine-rich domains (C); poly (A) tract (An). Within the L1 5’UTR monomer are highlighted: CpG island (CpG 1-13) and YY1 binding site (red), as reported in (31). **c)** Methylation analysis of L1MdTf promoter at YY1 binding site in hippocampus, cortex and cerebellum at different stages of embryonic (E15.5, E18.5) and postnatal development (P0, P3, P14). For each organ and developmental stage, samples from 4 different embryos/mice were analyzed. On an average 80,000 reads were analyzed for each sample. Hippocampus: E15.5 vs P14, p = 0.003; E18.5 vs P0, p = 0.047; E18.5 vs P3, p = 0.009; E18.5 vs P14, p = 0.000; P0 vs P14; p = 0.027. Cortex: E15.5 vs P14, p = 0.003; E18.5 vs P3, p = 0.031; E18.5 vs P14, p = 0.000; P0 vs P14, p = 0.001; P3 vs P14, p = 0.017, ANOVA with Tukey post hoc test. Data are represented as the mean percentage of methylation ± s.e.m.

We observed that L1MdTf promoter methylation at the YY1 binding site had a peculiar trend along hippocampus and cerebral cortex developmental trajectories, showing a wave of re-methylation at E18.5 followed by progressive demethylation postnatally (Figure 4c). To assess whether these dynamics were specific for L1 promoter, we analyzed IAPLTR1a (TEs belonging to ERV subfamilies) methylation as reported in (31) (see Methods), and found no changes in any of the sampled areas (Supplemental Figure 3a).

We next investigated whether the reduction in L1 methylation observed postnatally in mouse hippocampus and cortex could correspond to an increased L1 CNVs. We measured L1 CNVs in genomic DNA as reported in (42) (Figure 5a), treating the gDNA with Exonuclease I in order to avoid the amplification of L1 cDNA intermediates (31) (Supplemental Fig 3b). Interestingly, we observed a statistically significant increase in L1 CNVs specifically in hippocampus and cerebral cortex postnatally (P0, P3, P14), in correspondence with the decrease in L1 promoter methylation (Figure 5b).

**Figure 5.**
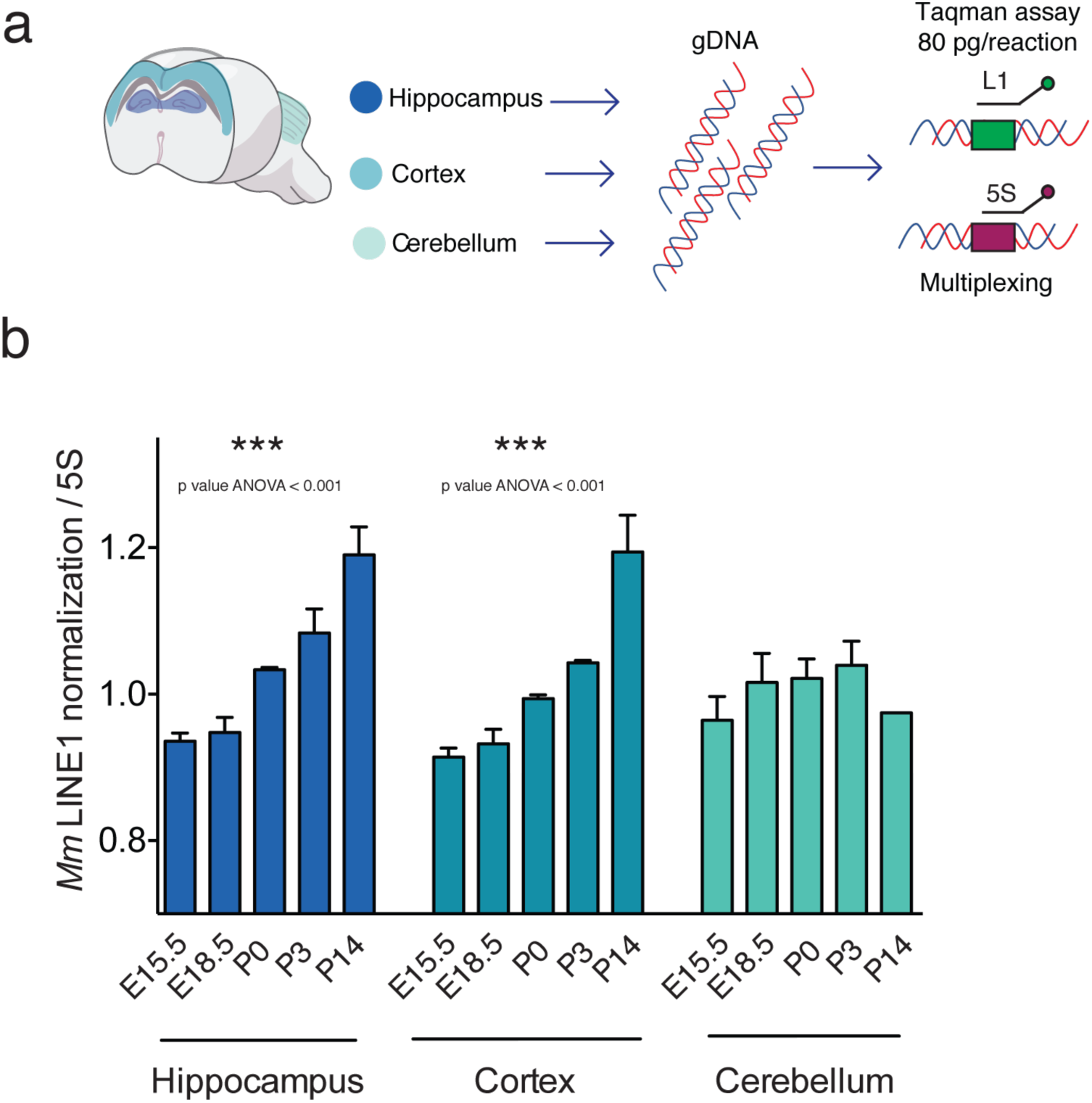
L1 CNVs early postnatally increase in mouse hippocampus and cortex. **a)** Schematic representation of L1 CNV assay performed on mouse hippocampus, cortex and cerebellum. Briefly, the assay is performed in multiplex qPCR, using Taqman probes specific for mL1-ORF2 and m5S as reported in (42). **b)** L1 CNV assay performed on mouse hippocampus, cortex and cerebellum at different stages of embryonic (E15.5, E18.5) and postnatal development (P0, P3, P14). For each organ and developmental stage, genomic DNA samples from the 4 mice (same as in Figure 4) were analyzed. mL1-ORF2 was normalized on m5S. Hippocampus: E15.5 vs P3, p = 0.006; E15.5 vs P14, p = 0.000; E18.5 vs P3, p = 0.012; E18.5 vs P14, p = 0.000; P0 vs P14, p = 0.004. Cortex: E15.5 vs P3, p = 0.018; E15.5 vs P14, p = 0.000; E18.5 vs P3, p =0.047; E18.5 vs P14, p = 0.000; P0 vs P14, p = 0.000, ANOVA with Tukey post hoc test. Data are represented as mean ± s.e.m.

Overall, these results suggest that L1 activity is fine-tuned during hippocampus and cortical development, and point at E18.5 as a sensitive and “vulnerable” developmental window for the epigenetic setting of L1 methylation and activity regulation in these brain regions.

### Early Intervention during NICU stay positively modulates neurodevelopmental outcomes in preterm infants

At 12 months corrected age, the preterm infants were tested with the Griffiths Mental Development Scales (GMDS) to assess their neurodevelopment (55). In the Standard Care group one infant could not participate to the follow-up, due to severe illness that required prolonged hospitalization after NICU discharge, and in the Early Intervention group one infant was excluded from the follow-up analysis, as he presented extensive white matter damage at brain MRI performed at 40^+0^ GA.

All the preterm infants showed scores within the normal range; however, statistically significant differences were observed between the 2 groups with the Early Intervention group showing higher scores (Table 2) in the General Quotient and in 4 out of 5 subscales: Personal-Social (that measures proficiency in the activities of daily living, level of independence and interaction with other children), Hearing and Speech (that assesses hearing, expressive language and receptive language), Eye and Hand Co-ordination (that tests fine motor skills, manual dexterity and visual monitoring skills) and Performance (that evaluates the ability to reason through tasks including speed of working and precision) (Table 2). No differences were observed in the Locomotor subscale that measures gross motor skills, including the ability to balance, coordinate and control movements (Table 2).

**Table 2.** Neurodevelopmental outcome at 12 months corrected age

| 12 Months Follow-up | Standard Care<br>(n = 15) | Early Intervention<br>(n = 17) | p value |
| --- | --- | --- | --- |
| General Quotient, mean ± SD | 90.5 ± 3.3 | 93.9 ± 4.4 | 0.017^ |
| Locomotor, mean ± SD | 96.3 ± 5.7 | 94.9 ± 9.8 | 0.894* |
| Personal-Social, mean ± SD | 87.5 ± 4.6 | 93.9 ± 5.1 | 0.001^ |
| Hearing and Speech, mean ± SD | 91.0 ± 4.2 | 94.8 ± 3.8 | 0.024* |
| Eye and Hand Coordination, mean ± SD | 89.5 ± 6.2 | 94.7 ± 4.8 | 0.014^ |
| Performance, mean ± SD | 91.1 ± 3.9 | 94.8 ± 5.6 | 0.026* |

Means ± standard deviations are shown. P-values were obtained using t-test (^) or Mann-Whitney U Test (\*).

## DISCUSSION

Here we report different L1 methylation patterns in preterm and full-term infants and we demonstrate that L1 methylation status in preterm infants can be modulated by the beneficial effect of maternal care and positive multisensory exposure. In addition, we demonstrate how the proposed early intervention strategy positively modulates short-term infants’ neurodevelopment. In this regard, it is already reported that maternal separation and excessive sensory exposure, induced by NICU environment, represent adverse early life events experienced by preterm infants, that can affect the epigenetic regulation and impact on gene expression in the brain (56-58).

Furthermore, previous studies have shown that L1 activity is modulated by early life experiences, reflecting on neuronal somatic mosaicism and genomic structural variations of the mouse hippocampus (30-34).

It is very well demonstrated that *de novo* L1 insertions are a developmentally regulated phenomenon that contributes to somatic mosaicism in the brain (59, 60). New L1 insertions could have an evolutionary adaptive significance, offering new genomic instruments to evolve into new phenotypes; an idea corroborated by the finding that new insertions land in neuronal specific active genes (37-39, 47). Moreover, L1 deregulated activity is now emerging as a potential mechanism that correlates with the development of several mental disorders including schizophrenia, autism spectrum disorders (ASD), major depression (43, 47, 50, 59, 61), which are also often described in former preterm infants (62). In this context, in the animal model we detected L1 CNVs as an early event (P0, P3) in hippocampus and cerebral cortex that match the progressive demethylation of L1 promoter after E18.5 stage. This suggests that in the mice E18.5 represents a susceptible time window in which L1 methylation status resets, soon before birth (Figure 4 and 5); it is tempting to speculate that similar dynamics could occur also in human brain development, although with a different timing.

Interestingly, hippocampus and the interconnected thalamus and cortex are known to be affected by preterm birth to an extent proportional to the degree of prematurity (13, 63) and to the premature exposure to the extrauterine environment (64, 65), highlighting how preterm birth disrupts specific aspects of cerebral development. Notably, these cerebral areas, in particular the hippocampus, are well-known for their involvement in socio-emotional development, functioning and memory (66). However, the relationship between the structural changes observed by neuroimaging studies in preterm infants, and the impairments in memory and learning that they manifest in childhood, are still debated (64) as well as the underlying pathogenetic mechanisms. No significant changes in L1 methylation and activity were observed in the cerebellum, although the third trimester of pregnancy is a highly dynamic period for cerebellar development which, in humans, follows a precisely programmed series of regionally- and temporally-specific (67) developmental processes (68). Different molecular mechanisms involved in these maturational processes or different time windows of vulnerability might explain this finding (67).

We described that L1 methylation levels are lower in preterm infants at birth and that they are properly restored only upon Early Intervention at NICU discharge, a recovery that specifically involve the YY1 binding site region. YY1 binding site methylation has been recently demonstrated to regulate L1 retrotransposition (52) and to be modulated by maternal care, (31) in turn altering L1 activity in mouse neural hippocampal genome (31). We can speculate that the specific alteration of L1 methylation that premature infants experience, and, more importantly, the inability to recover a proper L1 methylation setting could influence L1 activity, leading to aberrant L1 retrotransposition, as already demonstrated for several neurological disabilities (43, 47, 50, 59, 61).

We further demonstrate that the Early Intervention program enhances neurodevelopment at 12 months corrected age compared to Standard Care, albeit scores are within the normal range for both groups. The clinical relevance of these short-term results needs to be confirmed later in childhood when neurobehavioral and cognitive impairments become more obvious, although studies on the neurodevelopmental trajectories of preterm infants have suggested the predictive value of the 12 months’ assessment on long-term outcomes (69).

In conclusion, we are providing evidence that L1 methylation and activity are tightly developmentally controlled, being specifically influenced by the early life experiences of premature infants; this study sheds a new light on the epigenetic mechanisms related to a premature exposure to the extrauterine life and suggests that preterm birth affects the dynamic of L1 methylation. Early intervention strategies based on maternal care and positive sensory stimulation during a sensitive window of both epigenetic and brain plasticity might modulate L1 DNA methylation and contribute to shape the developing brain connections with potential impact on infants’ neurodevelopment.

## METHODS

### Study Cohort

All the preterm infants consecutively born between 25^+0^ and 29^+6^ weeks of gestational age (GA) at the same institution were eligible. Exclusion criteria: multiple pregnancy (triplets or higher), genetic syndromes and/or malformations, infants who developed severe neonatal comorbidities including severe brain lesions. The full study protocol is described in (51). Adherence to the early intervention protocol was required and documented in a parental self-report diary.

### Animals

CD-1 mice were housed under controlled conditions for temperature and humidity, using a 12:12-h light-dark cycle. Mice were mated overnight, and females were separated the following morning and checked for vaginal plugs (Embryonic day, E 0,5). CD-1 animals deliver pups between day E19 - E20. Caesarean sections (C-secs) were performed at embryonic days E15.5, E18.5 and P0. Pups were sacrificed by decapitation at different time points: at embryonic day E15.5, E18.5 and at postnatal day P0, P3 and P14. At each developmental stage, 4 mice were sacrificed and brains collected to manually microdissect hippocampal, cortical and cerebellar tissue under a stereomicroscope in sterile conditions. Microdissected tissues were store at −80°C until gDNA extraction was performed.

### Study design

Infants were randomly assigned either to receive (i) Standard Care or (ii) an additional Early Intervention protocol based on maternal care. Standard Care, according to the routine clinical protocol of the NICU, included Kangaroo Mother Care, minimal handling and non-pharmacological pain management. The Early Intervention protocol included, over routine clinical care, the PremieStart (70), which is based on parental involvement, and enriched multisensory stimulation proposed by parents after a period of training. This intervention included both tactile stimulation, through infant massage, performed twice a day and visual interaction provided once a day with a black and white toy or parents’ face. A complete detailed description of the intervention is available in (51).

The randomization was performed using sealed envelopes prepared in groups of 10 through computer-generated randomization. The randomization sequence was concealed until the group allocation was assigned, and the examiners (both biologist and psychologist that performed the follow-up examination) remained blinded for the entire study period.

The present study is part of a larger Randomized Controlled Trial (RCT) aimed at assessing the effectiveness of an Early Intervention program in promoting visual function and neurodevelopment in preterm infants. Within this context, exploratory analyses have been performed to investigate the effect of preterm birth and early interventions on L1 modulation.

### Sample Collection

In preterm infants, cord blood samples were collected at birth and peripheral blood samples were harvested at hospital discharge (around term equivalent age - TEA). Peripheral blood was obtained during blood sampling performed for routine blood examination, according to clinical practice. In healthy full-term infants’ cord blood samples were collected at birth only in infants born by C-section after uneventful pregnancies. Each sample consisted of 0.5 mL of cord/peripheral blood.

### Neurodevelopmental assessment

At 12 months corrected age the preterm infants group underwent the Griffiths Mental Development Scales (GMDS) to asses neurodevelopment (71). This evaluation comprises five subscales (score range 50–150): Locomotor, Personal-Social, Hearing and Speech, Eye and Hand Coordination and Performance. The subscales yield standardized scores for each domain (mean ± SD: 100 ± 16) and a composite General Quotient (mean ± SD: 100 ± 12).

### DNA extraction

Genomic DNA from human cord and peripheral blood, from mouse hippocampus, cortex and cerebellum was isolated with standard phenol-chloroform extraction techniques.

### Bisulfite conversion

500 ng of genomic DNA from each sample were bisulfite-treated using the MethylEdge™ Bisulfite Conversion System (Promega, Madison, USA) following the manufacturer’s protocol.

### Methylation assay in human samples

The methylation analysis of CpG island within the human L1 promoter were conducted as reported in (42) with minor modifications. The primer sequences are the following:

hL1-5’UTR For: 5’ - AAGGGGTTAGGGAGTTTTTTT – 3’

hL1-5’UTR Rev: 5’ - TATCTATACCCTACCCCCAAAA – 3’

In each PCR, 40 ng of bisulfite-converted DNA were combined with primers at 0.5 μM final concentration and GoTaq™ Hot Start Green Master Mix (Promega) in a final volume of 50 μL. PCR conditions were as follows: 95°C for 2 minutes followed by 30 cycles of 95 °C for 45 seconds, 56 °C for 1 minute and 72°C for 30 seconds, followed by a final step of 72°C hold for 4 minutes.

The product of amplification is 363 bp of length and contains 19 CpGs. The resulting PCR products were checked by agarose gel electrophoresis and then purified by PureLink ™ Quick Gel Extraction & PCR Purification Combo Kit (Invitrogen-Thermo Fisher Scientific, USA). They were then cloned into pGEM-T Easy Vector System I (Promega) using a molar ratio insert: vector of 6:1. Sanger sequencing was performed by GATC Biotech, using the reverse sequencing primer pGEM Seq Rev: 5’-GACCATGATTACGCCAAGCTA – 3’. Resulting chromatograms were examined for sequencing quality using FinchTV software. At least 10 sequenced clones per sample were analyzed for Figure 3 and Supplemental Figure 1 and at least 11 clones per sample were analyzed in Supplemental Figure 2b and c as suggested in (72).

The methylation analysis of CpG island within the *Nab2* promoter region were conducted designing specific primers with MethPrimer (73), whose sequences are as follows

*Nab2* For: 5’ - GAGGGAGGGATAGAGTTTGGAT - 3’

*Nab2* Rev: 5’ - ACGCTCTATACATAAACGACCGA - 3’

In each PCR, 80ng of bisulfite-converted DNA were combined with primers at 0.5 μM final concentration and GoTaq™ Hot Start Green Master Mix (Promega), in a final volume of 50 μL. PCR conditions were as follows: 95°C for 4 minutes followed by 35 cycles of 95 °C for 45 seconds, 58 °C for 1:30 minutes and 72°C for 2 minutes, followed by a final step of 72°C hold for 4 minutes.

The product of amplification is 135-bp amplicon and contains 15 CpGs. The resulting PCR products were checked by agarose gel electrophoresis and then purified by PureLink ™ Quick Gel Extraction & PCR Purification Combo Kit (Invitrogen-Thermo Fisher Scientific, USA). Once purified, they were cloned into pGEM-T Easy Vector System I (Promega) using a molar ratio insert: vector of 6:1. Sanger sequencing was performed by GATC Biotech, using the reverse sequencing primer pGEM Seq For: 5’ - ACGACGGCCAGTGAATTG – 3’. Resulting chromatograms were examined for sequencing quality using FinchTV software. At least 5 sequenced clones per sample were analyzed as suggested in (72).

### Analysis of Sanger sequencing in human samples

To analyze the conversion efficiency and the methylation status of the CpG sites, FASTAQ files were analyzed by QUMA (*<u>QU</u>antification tool for <u>M</u>ethylation <u>A</u>nalysis*) software (CDB, Riken, Japan) (74). For the L1 promoter methylation, we excluded from the analysis three (CpG 2, 6 and 9) of the 19 CpGs due to the high degree of variability among the analyzed sequences compared to the consensus sequence used (L19092.1 Human LINE1 (L1.4)). Sequences with a >90% of cytosine residues converted were used for subsequent analisys. Total percent methylation was calculated as the number of methylated CpGs divided by the number of total CpGs (both methylated and unmethylated) multiplied by 100. To determine the methylation status of each CpG site, we calculated the percentage of methylation of each CpG site as the number of methylation events at a specific CpG site divided by the total number of sequenced and analyzed clones.

### Methylation assay in mouse samples

The methylation analysis of CpG island within the murine L1MdTf monomer and IAPLTR1a were conducted as reported in (31) with minor modifications. Given the peculiar monomeric and highly repeated nature of the mouse L15’UTR we performed this methylation analysis with a Next Generation sequences approach. A detailed list of primer sequences used for the amplification is reported in Supplemental Table I. Briefly both for L1MdTf monomer and IAPLTR1a we used primers with Illumina barcode index (Illumina Truseq LT 6-mer indices): each organ in each developmental stage was associated to a distinct couple of Fow and Rev primers 5’ end tagged, in order to be unambiguously identified in the sequencing analysis step (see Supplemental Table I).

Each PCR was performed with 16 ng – 40 ng of bisulfite-converted DNA were combined with primers at 0.5 μM final concentration and GoTaq™ Hot Start Green Master Mix (Promega) in a final volume of 50 μL. PCR conditions were as follows: 95°C for 2 minutes followed by 30 cycles of 95 °C for 45 seconds; 56 °C for 1 minute and 72°C for 5 seconds, followed by a final step of 72°C hold for 4 minutes. For L1MdTf monomer the product of amplification is 191-bp of length and contains 13 CpGs while for IAPLTR1a the product of amplification is 205 bp of length and contains 10 CpGs. The resulting PCR products were checked by agarose gel electrophoresis and then purified by Agencourt AMPure XP beads (Beckman Coulter) according to the manufacturer’s instructions. DNA concentration were quantified using a Qubit dsDNA HS Assay kit. All the L1MdTf and IAPLTR1a amplicons were then pooled in equimolar quantities to obtain a final pooling concentration of 2 ng/μL. Library for DNA sequencing was produced on the pooled PCRs. Paired-end 2 x 150bp sequencing was performed on a HiSeq platform (Illumina) by Eurofins GATC Biotech.

### Analysis of NGS sequencing in mouse samples

A total of 22,481,000 reads were obtained from bisulfite sequencing and were assigned to samples based on the primers with Illumina barcode index. Briefly, no mismatch was allowed for the barcode index and a maximum of 5 mismatches were allowed for the target primer. None of the reads assigned to IAPLTR1a target aligned on L1MdTf and vice versa. Prior to mapping, reads were trimmed for low quality using Trimmomatic (75) (parameters: ILLUMINACLIP:TruSeq3-PE.fa:2:30:10 LEADING:3 TRAILING:3 SLIDINGWINDOW:4:15 MINLEN:75). 15,368,000 reads were obtained post trimming with an average of 80,000 reads for each sample. The paired reads were mapped using Bismark (76) (parameters: --local -N 1 -L 15 --non_directional) with an average mapping efficiency 99.05%. DNA methylation data was called using MethylDackel (https://github.com/dpryan79/MethylDackel). Sample correlation analysis was performed using methylKit (77) and all biological replicates of a given organ within a developmental stage showed a correlation higher than 95%. Methylation analysis for L1MdTf was focused on the YY1 binding site, corresponding to the CpG sites 8 and 9, as reported by (31) and on four CpG sites for IAPLTR1a, as reported by (31). Briefly, methylation status of each CpG site, was calculated as the number of methylation events at a specific CpG site divided by the total number of analyzed sequences. Sample methylation level was calculated as the number of methylated CpGs divided by the number of total CpGs (both methylated and unmethylated) multiplied by 100.

### TaqMan PCR for L1 expression and CNVs analysis

For L1 CNVs 300 ng of genomic DNA was treated with Exonuclease I, following manufacturer instructions (40U of Exonuclease I, in 1X Reaction Buffer 67 mM glycine-KOH (pH 9.5 at 25 °C), 6,7 mM MgCl2, 1 mM DTT were used at 37°C for 30 min, then inactivated at 85°C for 15 min). Efficiency of digestion was proved on 300ng of gDNA equimolar pooled with 300 ng of a 120 bp ssDNA oligonucleotide (Supplemental Figure 3c). Digested DNA was further subjected to phenol-chloroform purification. Extracted DNA was quantified using Qbit HS DNA kit (Invitrogen) and diluted to a concentration of 80 pg/μL.

Quantitative PCR experiments were performed on a StepOne Plus (Thermo Fisher Scientific) with minor modifications to the method reported in (42). In each multiplexed PCR two TaqMan probes, labeled FAM and VIC, were combined; 80 pg of genomic DNA was combined with gene-specific primers, TaqMan-MGB probes and 10 μL of iQ multiplex PowerMix (Biorad) in a total volume of 20 μL. Primers’ concentration was 0.4 μM and TaqMan probes’ concentration 0.4 μM. PCR conditions were as follows: 95°C for 2 minutes followed by 40 cycles of 95°C for 10 seconds and 59°C for 60 seconds.

Standard curves of genomic DNA ranging from 200 ng to 0.2 ng were performed to verify that the 80 pg dilution was within the linear range of the reaction. For CNVs, the quantification includes from five to eight technical replicates. For assays on mouse genome, we adapted a TaqMan probe for the same amplicon reported in (31, 43). Probes’ and primers’ sequences are reported below:

mL1 ORF2 F: 5’ – CTGGCGAGGATGTGGAGAA - 3’

mL1 ORF2 R: 5’ – CCTGCAATCCCACCAACAT - 3’

mL1 ORF2 Taqman probe: 5’ – TGGAGAAAGAGGAACACTCCTCC - 3’

mL1 5S F: 5’ – ACGGCCATACCACCCTGAAC - 3’

mL1 5S R: 5’ – AGCCTACAGCACCCGGTATTC - 3’

mL1 5S Taqman probe: 5’ – GATCTCGTCTGATCTCGGAAGCTAAG - 3’

### FACS analysis and isolation of PBMCs populations

Fresh cord blood samples derived from full-term and preterm were subjected to erythrocytes lysis following manufacturer’s instruction (BD lysis buffer). PBMCs (Peripheral Blood Mononuclear Cells) were then stained with anti CD45 for 30 minutes at 37°C, different mononuclear subpopulations were identified gating on CD45 and SSC as described in (78). Most abundant populations as granulocytes and lymphocytes were then sorted to be further subjected to DNA methylation analysis. granulocytes were sorted as the population CD45 high with the highest SSC while lymphocytes were sorted as the population CD45 high with the lowest SSC.

### Study approval

The present study was approved by the Ethics Committee Milano Area B. The trial is registered at Clinicaltrial.gov (NCT02983513). Written informed consent was signed by both parents before inclusion in the study (both for preterm and full-term infants). All experimental procedures were performed in compliance with national and EU legislation, and Humanitas Clinical and Research Center, approved by Animal Care and Use Committee.

### Accession number

The data from bisulfite sequencing has been submitted in NCBI GEO (GSE136844).

## STATISTICAL METHODS

Demographic and baseline characteristics were described as mean ± SD, median and range or number and percentage. Independent t-test and Mann-Whitney U test were used in the comparison of continuous variables with normal distribution and non-normal distribution respectively. For the comparison of qualitative data, Fisher’s exact test was used. Shapiro-Wilk test was used to test the normal distribution of the data. To assess the differences between full-term and preterm infants, and between treatment groups in total L1 methylation and on each CpG, unpaired t-test and two-way ANOVA model with Tukey’s HSD post-hoc tests were used. Linear regression model was used to study the relationship between L1 methylation at NICU discharge and the intensity of care (mean number of massages per week) and independent t-test and Mann-Whitney U test were used to assess the difference in 12 months neurodevelopment between Standard Care and Early Intervention groups.

Mouse brain regions’ methylation at different stages of development were analyzed using one-way ANOVA and Tukey’s HSD post-hoc tests. All tests were two-tailed, and p < 0.05 was considered significant for all tests. Statistical analyses were performed using R version 3.5.3 (R Foundation for Statistical Computing, Vienna, Austria).

## Data Availability

Data available upon request.

## AUTHOR CONTRIBUTIONS

C.F. designed and performed clinical developmental intervention, analyzed the data and wrote the manuscript F.M. designed and performed molecular experiments, analyzed the data and wrote the manuscript. L.P. performed molecular experiments and wrote the manuscript. S.M. isolated and collected mouse brain areas. N.P. performed statistical analyses. S.S. performed bioinformatical analyses. S.P. contributed to data collection and interpretation. S.A. and F.M. contributed to study design and data interpretation. S.L. supervised mouse data collection, contributed to data interpretation and write the manuscript. B.B. and M.F. conceived this study, designed experiments, analyzed the data and wrote the manuscript.

## AKNOWLEDGMENTS

We thank C. Lanzuolo for the stimulating discussions and constructive criticisms of this manuscript. The authors acknowledge the technical assistance of the INGM Sorting Facility (Istituto Nazionale di Genetica Molecolare “Romeo ed Enrica Invernizzi” (INGM), Milan, Italy), in particular M.C. Crosti. We are grateful to the infants and families who participated in the study. We thank C. Lembo, the staff of the NICU and of the preterms’ follow-up clinic (in particular O. Picciolini, S. Gangi, L. Gardon and C. Squarza), for supporting the project.

We acknowledge funding from the Italian Ministry of Health (RC 780/03 2017), the University of Milan (DISCCO 2015) and INGM internal funding

